# Evaluating agentic simulation for local public health modelling

**DOI:** 10.64898/2026.09.19.26363431

**Authors:** Michelle A. Nakatsuka, Robert J. Steele, Xu Han, Eric K. Oermann

## Abstract

Large language model (LLM)-based generative agents can reproduce aspects of individual human behavior, but whether they can be scaled to geographically grounded populations that reproduce real-world health behaviors remains unclear. Here, we introduce LLMPopSim, an agentic population simulation framework that uses U.S. Census and Centers for Disease Control and Prevention data to construct populations of AI agents whose simulated individual health behaviors can be aggregated and evaluated at the community level. We developed the framework using historical data from Hawaiʻi in 2018, and evaluated temporal and geographic generalizability using held-out 2022 cohorts from Hawaiʻi and New York State, with colorectal cancer screening and mammography as proof-of-concept behaviors. Across the four state-outcome evaluations, mean absolute error ranged from 3.5 to 15.0 percentage points and correlations between simulated and observed ZCTA-level prevalence ranged from 0.26 to 0.69. Mean prediction bias was +15.0 and +10.2 percentage points for colorectal cancer screening in Hawaiʻi and New York State, respectively, and +2.9 and +0.9 percentage points for mammography; ratios of predicted to observed geographic standard deviation were 0.60 and 0.68 for colorectal cancer screening and 1.12 and 1.96 for mammography, respectively. Prediction error was greatest in communities with lower observed screening prevalence and varied across community characteristics without a uniform socioeconomic gradient. These findings demonstrate that individually represented AI agents can aggregate into population-level patterns that retain measurable features of real-world health behavior across temporal and geographic transfer, while identifying calibration, distributional fidelity and subgroup performance as key challenges for generative population simulation. LLMPopSim provides an empirical foundation for developing synthetic populations that may ultimately enable simulation of heterogeneous population responses to public health interventions.

## Introduction

Understanding how health behaviors emerge across populations is fundamental to public health research, health policy, and intervention design.^1,2^ Population-level models provide a means to characterize geographic variation in health behaviors, identify communities at increased risk, and evaluate strategies for improving healthcare delivery and reducing disparities. Preventive behaviors such as colorectal cancer screening and mammography vary substantially across communities and are influenced by socioeconomic conditions, race and ethnicity, healthcare access, and local infrastructure.^3-5^ Because these patterns often occur at scales smaller than counties or states, geographically resolved population models have become increasingly important for understanding localized health disparities and informing targeted public health interventions.^1,2^

Current approaches to population health modeling primarily rely on survey systems such as the Behavioral Risk Factor Surveillance System (BRFSS) coupled with statistical small-area estimation frameworks including multilevel regression and poststratification.^6,7^ These approaches have enabled geographically resolved surveillance platforms such as the Centers for Disease Control and Prevention (CDC) PLACES project.^2^ However, they are fundamentally designed to estimate observed outcomes rather than simulate populations or explore the mechanisms that give rise to those outcomes. Because direct survey observations are often sparse at fine geographic scales, small-area estimation methods rely on statistical models that combine survey data with demographic and geographic information to generate local estimates.^2,6,7^ Alternative data sources, including electronic health records, insurance claims, and digital trace data, provide complementary information but introduce their own challenges related to representativeness, accessibility, and systematic bias.^8,9^ Together, these limitations highlight the need for complementary approaches capable of generating realistic synthetic populations while preserving meaningful geographic variation in health behaviors.

Recent advances in large language models (LLMs) and agentic simulation suggest a fundamentally different paradigm for computational population health. LLM-based agents can be instantiated with individual characteristics and contextual information to generate behavior within simulated environments, and emerging work suggests that these agents can reproduce aspects of human reasoning, preferences, and behavior, ranging from emergent social interactions among synthetic agents to the replication of survey responses and experimental behaviors of real individuals.^10,11^ These findings raise the possibility that LLMs could serve not only as language generation systems, but also as engines for generative population simulation, enabling the construction of synthetic populations whose emergent behaviors can be evaluated against those of real-world communities.

Despite growing interest in generative agents, applications to population-level public health remain nascent. Recent studies have begun to construct demographically grounded agent populations to simulate health-related decisions and responses to public health interventions, including vaccine uptake and mobility responses during infectious disease outbreaks.^12,13^ However, important questions remain regarding the empirical calibration of LLM-generated population behaviors, their ability to preserve fine-scale geographic heterogeneity, and their performance across communities with differing demographic and socioeconomic characteristics. Addressing these questions is essential for establishing the conditions under which LLM-based population simulation may provide a reliable framework for public health research, surveillance, and policy evaluation.

Here, we present LLMPopSim, a generative population simulation framework that combines Census-derived synthetic populations, CDC PLACES data, and LLM-based behavioral simulation to model preventive health behaviors at the ZIP Code Tabulation Area (ZCTA) level. We developed and optimized the framework using multiple years of Hawaiʻi data and subsequently evaluated its performance and geographic generalizability using held-out 2022 data from Hawaiʻi and New York State. As an initial proof of concept, we evaluated whether LLMPopSim could reproduce community-level colorectal cancer screening and mammography prevalence while preserving geographic heterogeneity and demographic subgroup patterns. Together, these analyses establish small-area estimation as an initial application of a broader LLM-driven population simulation framework and provide an empirical evaluation of its potential for geographically grounded public health modeling (Figure 1).

**Figure 1.**
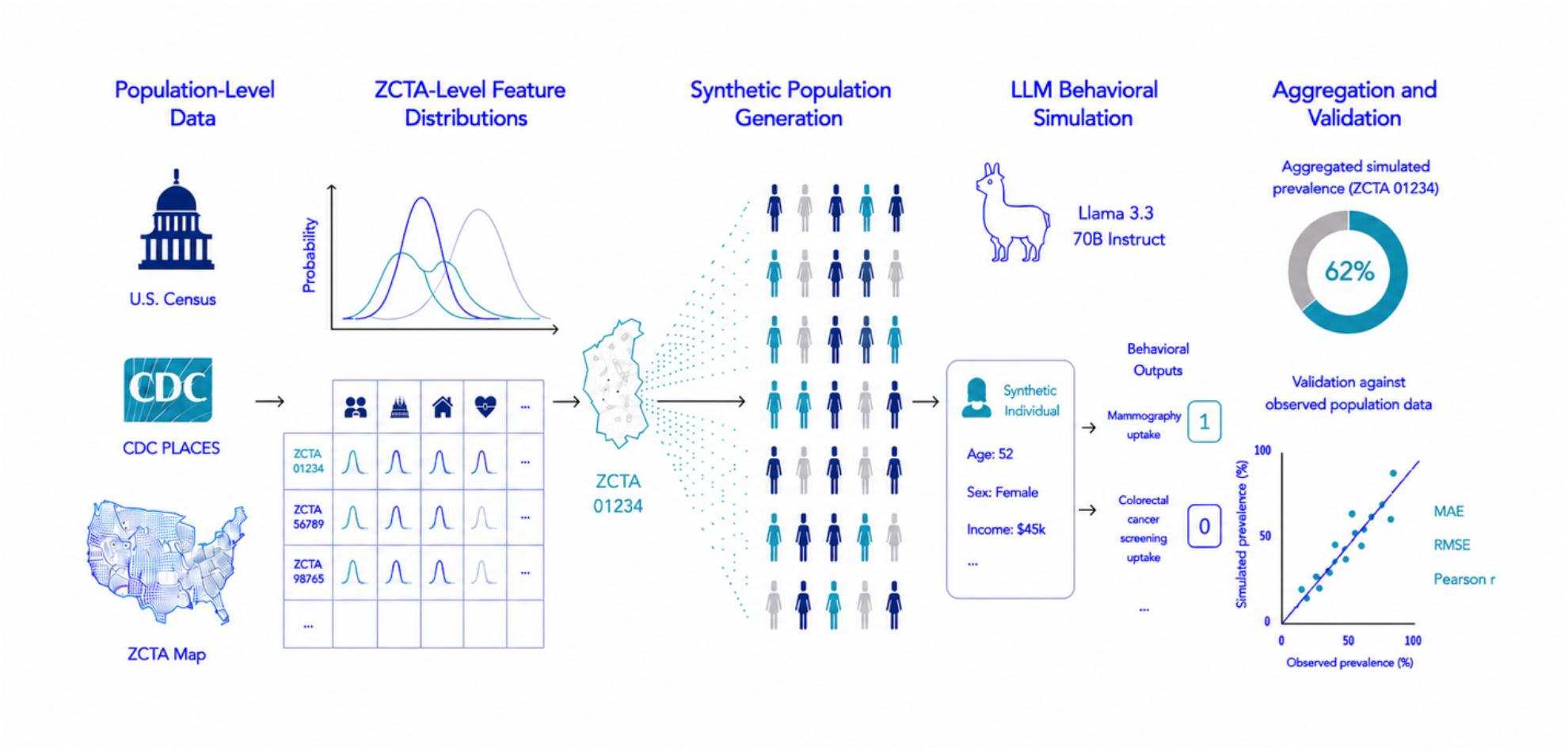
LLMPopSim framework for geographically grounded generative population simulation. Population-level demographic, socioeconomic and health data from the U.S. Census Bureau and CDC PLACES were integrated to construct ZCTA-specific feature distributions. Synthetic individuals were probabilistically sampled to generate geographically grounded populations representing each ZCTA. Structured individual-level profiles were provided to an instruction-tuned large language model to simulate preventive health behaviors. Individual responses were aggregated to estimate community-level prevalence and evaluated against CDC PLACES estimates. Framework development was conducted using historical Hawaiʻi data, followed by held-out evaluation in 2022 Hawaiʻi and New York State cohorts.

## Results

### Performance in held-out temporal and geographic test cohorts

LLMPopSim was evaluated using held-out 2022 data from Hawaiʻi and New York State, comprising 94 and 1,745 ZCTAs, respectively. In Hawaiʻi, colorectal cancer (CRC) screening predictions were available for 94 ZCTAs. Mean predicted prevalence was 77.8%, compared with a mean observed prevalence of 62.8%, corresponding to a mean absolute error (MAE) of 15.0 percentage points (pp), root mean squared error (RMSE) of 15.5 pp, and mean bias of +15.0 pp. Despite this systematic overestimation, predicted prevalence was positively correlated with observed geographic variation across ZCTAs (Pearson r = 0.69; Fig. 2a; Table 1). For mammography, predictions were available for 93 ZCTAs. Mean predicted prevalence was 77.8%, compared with a mean observed prevalence of 75.0%, with an MAE of 4.15 pp, RMSE of 5.36 pp, and mean bias of +2.85 pp. The correlation between predicted and observed mammography prevalence was weaker (r = 0.26; Fig. 2b; Table 1).

**Table 1.** Overall performance of LLMPopSim in held-out 2022 cohorts. Across the four state-outcome combinations, MAE ranged from 3.5 to 15.0 percentage points, RMSE from 4.9 to 15.5 percentage points and Pearson correlations from 0.26 to 0.69. Performance varied by outcome and geography, with differences between absolute predictive error, geographic correlation and calibration.

| Metric | Hawai'i CRC | Hawai'i Mammography | New York CRC | New York Mammography |
| --- | --- | --- | --- | --- |
| ZCTAs, n | 94 | 93 | 1,744 | 1,741 |
| Mean observed prevalence, % | 62.85 | 74.96 | 69.23 | 78.27 |
| Mean predicted prevalence, % | 77.84 | 77.81 | 79.43 | 79.12 |
| Mean bias, pp | +14.99 | +2.85 | +10.20 | +0.85 |
| MAE, pp | 14.99 | 4.15 | 10.23 | 3.49 |
| RMSE, pp | 15.50 | 5.36 | 10.86 | 4.88 |
| Pearson $r$ | 0.69 | 0.26 | 0.59 | 0.40 |
| $R^2$ | 0.48 | 0.07 | 0.35 | 0.16 |
| Calibration intercept | -26.08 | 56.44 | 0.42 | 62.20 |
| Calibration slope | 1.14 | 0.24 | 0.87 | 0.20 |
| Observed SD, pp | 5.5 | 3.6 | 4.6 | 2.6 |
| Predicted SD, pp | 3.3 | 4.0 | 3.1 | 5.2 |
| Predicted/observed SD ratio | 0.60 | 1.12 | 0.68 | 1.96 |

**Figure 2.**
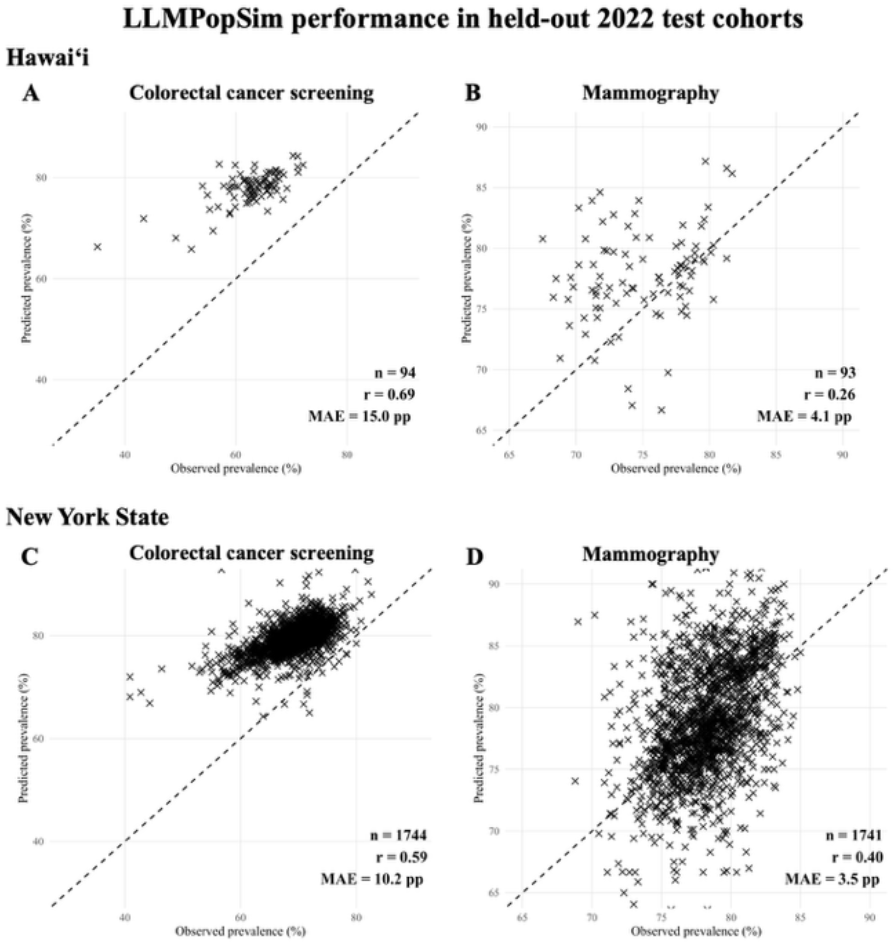
LLMPopSim reproduces ZCTA-level preventive screening patterns across Hawaiʻi and New York State. Observed CDC PLACES prevalence is plotted against LLMPopSim-predicted prevalence for (A) colorectal cancer screening in Hawaiʻi, (B) mammography in Hawaiʻi, (C) colorectal cancer screening in New York State and (D) mammography in New York State. Each point represents one ZCTA. Dashed lines indicate perfect agreement. Insets report the number of ZCTAs, Pearson correlation coefficient and mean absolute error in percentage points.

Geographic generalization was subsequently evaluated in New York State. Among 1,744 ZCTAs with complete CRC screening estimates, mean predicted prevalence was 79.4%, compared with a mean observed prevalence of 69.2%, yielding an MAE of 10.2 pp, RMSE of 10.9 pp, and mean bias of +10.2 pp. Predicted and observed CRC prevalence were moderately correlated across ZCTAs (r = 0.59; Fig. 2c; Table 1). Mammography predictions were available for 1,741 ZCTAs and demonstrated closer agreement in absolute prevalence: mean predicted and observed prevalences were 79.1% and 78.3%, respectively, with an MAE of 3.49 pp, RMSE of 4.88 pp, and mean bias of +0.85 pp. Predicted and observed mammography prevalence were correlated at r = 0.40 (Fig. 2d; Table 1).

### Preservation of geographic variation in screening prevalence

The extent to which simulated estimates reproduced the geographic dispersion of observed screening prevalence differed by outcome and setting (Fig. 3). In Hawaiʻi, the standard deviation (SD) of predicted CRC prevalence across ZCTAs was 3.3 pp, compared with 5.5 pp for observed prevalence, corresponding to a predicted-to-observed SD ratio of 0.60 (Fig. 3a). In contrast, the predicted mammography distribution more closely reproduced the observed dispersion, with predicted and observed SDs of 4.0 and 3.6 pp, respectively (SD ratio, 1.12; Fig. 3b).

**Figure 3.**
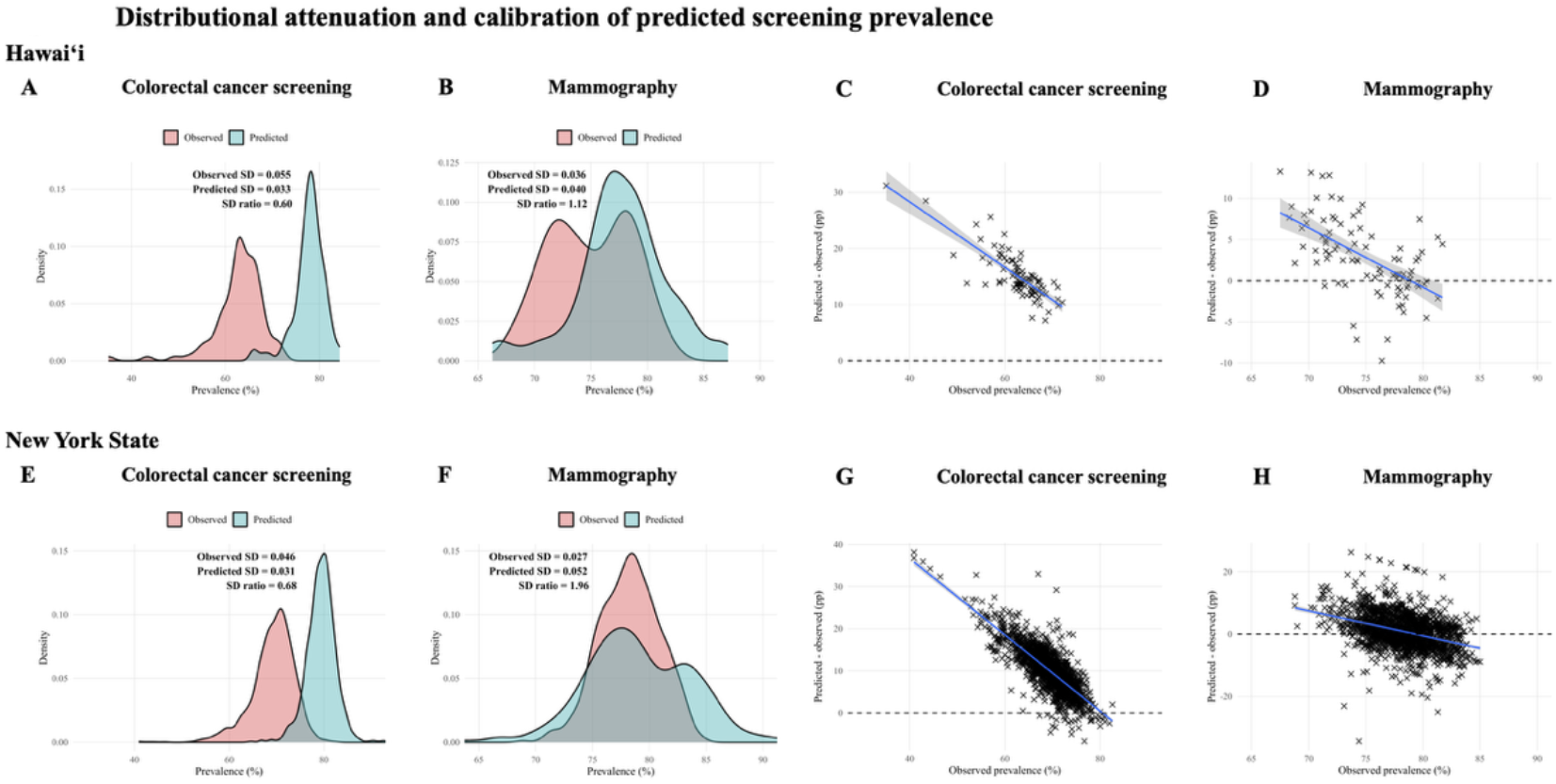
Outcome-dependent distortion of geographic prevalence distributions. Observed and LLMPopSim-predicted prevalence distributions are shown for (A) colorectal cancer screening and (B) mammography in Hawaiʻi and (E) colorectal cancer screening and (F) mammography in New York State. Insets report the observed and predicted standard deviations and their ratio, with values below 1 indicating variance compression and values above 1 indicating variance expansion. Residual error, defined as predicted minus observed prevalence, is plotted against observed prevalence for (C) colorectal cancer screening and (D) mammography in Hawaiʻi and (G) colorectal cancer screening and (H) mammography in New York State. Solid lines indicate linear regression fits with shaded 95% confidence intervals; dashed horizontal lines indicate zero residual error.

Residual analyses in Hawaiʻi further demonstrated outcome-specific calibration patterns (Fig. 3c, Fig. 3d). CRC predictions were predominantly higher than observed prevalence, whereas mammography residuals were centered closer to zero, particularly among ZCTAs with higher observed mammography prevalence.

A similar reduction in CRC variability was observed in New York State, where predicted and observed SDs were 3.1 and 4.6 pp, respectively (SD ratio, 0.68; Fig. 3e). Mammography showed the opposite pattern: predicted prevalence varied more widely across New York ZCTAs than observed prevalence, with predicted and observed SDs of 5.2 and 2.6 pp, respectively (SD ratio, 1.96; Fig. 3f). Consistent with these distributional differences, calibration slopes for CRC and mammography were 1.14 and 0.24, respectively, in Hawaiʻi and 0.87 and 0.20, respectively, in New York State. Residual analyses in New York State showed a similar outcome-specific pattern (Fig. 3g, Fig. 3h), with CRC predictions predominantly higher than observed prevalence and mammography residuals centered closer to zero (Table 1).

### Prediction error across the observed prevalence

Prediction error varied systematically across the observed screening prevalence distribution in both test settings (Fig. 4c, Fig. 4f). In Hawaiʻi, mean CRC residual error decreased monotonically across increasing quartiles of observed prevalence, from +19.4 pp in the lowest prevalence quartile to +15.0 pp, +13.5 pp, and +11.9 pp in successive quartiles. A similar gradient was observed in New York State, where mean CRC residual error decreased from +13.8 pp in the lowest prevalence quartile to +10.6 pp, +9.2 pp, and +7.3 pp across increasing prevalence quartiles. CRC absolute error followed the same pattern, with the largest discrepancies occurring among ZCTAs with the lowest observed CRC screening prevalence.

**Figure 4.**
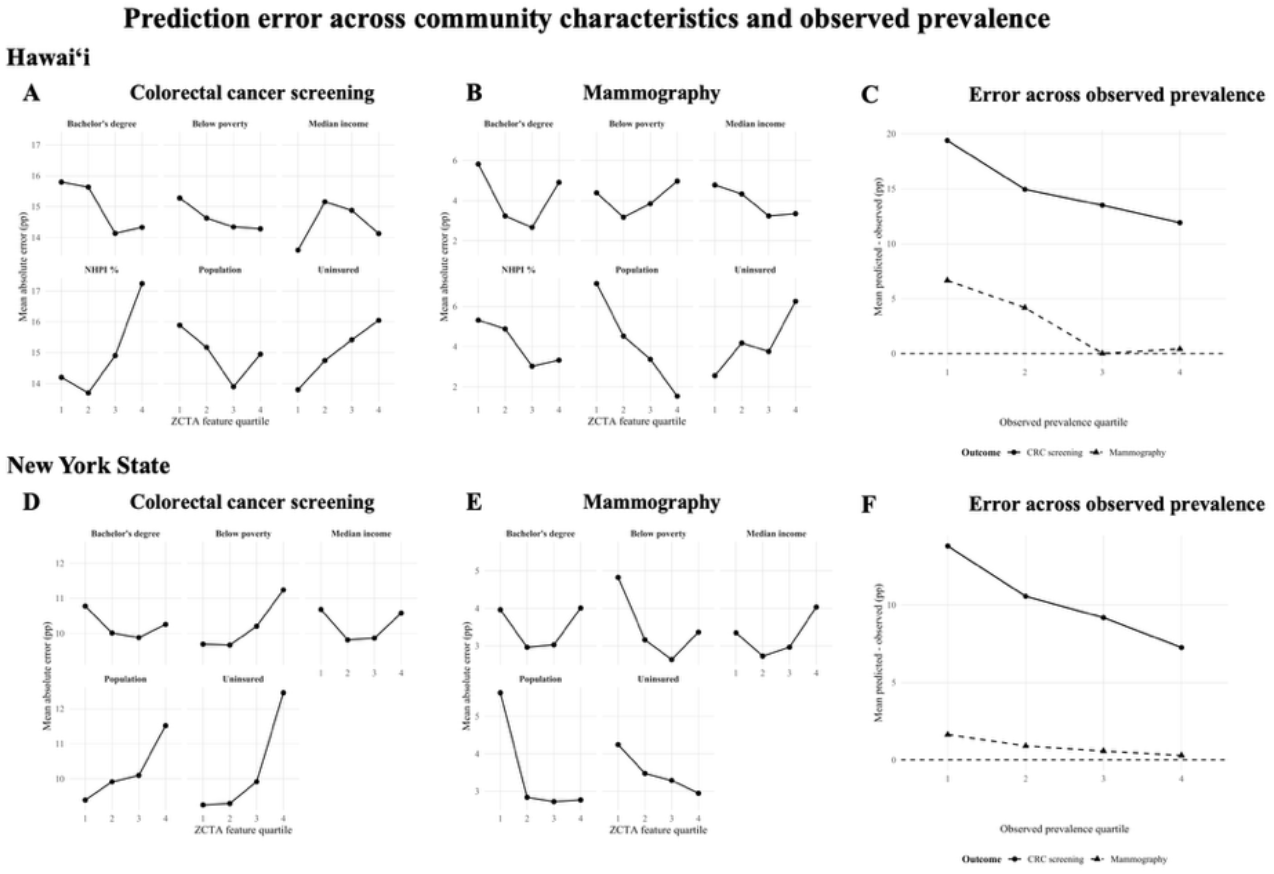
LLMPopSim performance across community characteristics and observed prevalence. Mean absolute error for (A) colorectal cancer screening and (B) mammography is shown across quartiles of selected ZCTA-level community characteristics in Hawaiʻi, with (C) mean prediction bias across quartiles of observed screening prevalence. Corresponding analyses are shown for (D) colorectal cancer screening, (E) mammography and (F) observed prevalence quartiles in New York State. Community characteristics included median household income, population size, poverty, bachelor’s degree attainment and PLACES-estimated uninsured prevalence among adults aged 18-64; Native Hawaiian and Pacific Islander population share was additionally evaluated in Hawaiʻi. Quartiles were defined separately within each state. Prediction bias was defined as predicted minus observed prevalence.

Mammography demonstrated smaller but directionally similar differences. In Hawaiʻi, mean residual error decreased from +6.65 pp in the lowest observed prevalence quartile to +4.17 pp in the second quartile and was near zero in the upper two quartiles (+0.01 and +0.41 pp, respectively). Mammography MAE correspondingly decreased from 6.70 pp in the lowest prevalence quartile to 2.12 pp in the highest. In New York State, mean mammography residual error decreased from +1.63 pp in the lowest prevalence quartile to +0.30 pp in the highest, while MAE remained comparatively stable across quartiles (3.25-3.88 pp). Across both geographic settings and screening outcomes, positive prediction bias was therefore greatest among ZCTAs with the lowest observed screening prevalence.

### Performance across community characteristics

Prediction error also varied across selected ZCTA demographic and socioeconomic characteristics, although patterns differed by screening outcome and geographic setting (Fig. 4a, Fig. 4b). In Hawaiʻi, CRC MAE was relatively stable across quartiles of median income, poverty, educational attainment, and population size. Across quartiles of Native Hawaiian and Pacific Islander (NHPI) population composition, CRC MAE was 14.2, 13.7, 14.9, and 17.2 pp, respectively, with the greatest error observed among ZCTAs in the highest NHPI-composition quartile. This pattern was not observed for mammography, for which MAE across increasing NHPI-composition quartiles was 5.32, 4.88, 3.03, and 3.33 pp. Mammography error varied more prominently by population size, decreasing from 7.14 pp in the lowest population quartile to 1.54 pp in the highest. Mammography MAE was also higher in the highest compared with the lowest quartile of uninsured prevalence (6.26 versus 2.55 pp).

In New York State, CRC MAE increased across quartiles of uninsured prevalence, from 9.25 pp in the lowest quartile to 12.46 pp in the highest, and across poverty quartiles, from 9.69 to 11.24 pp. CRC MAE also increased with ZCTA population size, from 9.39 pp in the lowest population quartile to 11.52 pp in the highest, whereas differences across median income and bachelor’s degree quartiles were smaller. Mammography demonstrated a different pattern: MAE was greatest among ZCTAs in the lowest population quartile (5.63 pp) and ranged from 2.73 to 2.84 pp across the remaining population quartiles. Error across income and educational attainment quartiles was non-monotonic, while mammography MAE decreased from 4.24 pp in the lowest uninsured quartile to 2.95 pp in the highest. Overall, subgroup patterns differed by screening outcome and geographic setting rather than demonstrating a uniform socioeconomic gradient in prediction error (Fig. 4d, Fig. 4e).

## Discussion

Here, we demonstrate that generative agents can be extended from simulations of individual behavior to geographically grounded synthetic populations that recover measurable features of real-world population health behavior. LLMPopSim generated community-level estimates of colorectal cancer screening and mammography prevalence that captured geographic variation in temporally held-out Hawaiʻi data and, importantly, after transfer to a geographically distinct population in New York State. However, this correspondence was incomplete and varied substantially across outcomes and dimensions of model performance. Geographic correlation did not necessarily coincide with accurate calibration or faithful reproduction of between-community variation, and prediction error varied across the underlying prevalence distribution and community characteristics. Together, these findings provide proof of concept for empirically grounded LLM-based population simulation while identifying key dimensions of population fidelity that must be considered as these systems are developed for broader public health applications.

A central finding of this study is that fidelity in generative population simulation is multidimensional. Measures of absolute error, geographic correlation, calibration and distributional fidelity frequently diverged across evaluations, indicating that no single performance metric adequately characterized how well the simulated populations reproduced observed community-level behavior. This was particularly evident for colorectal cancer screening, for which LLMPopSim preserved geographic ranking relatively well across both states despite systematic overestimation and compression of between-community variation. Mammography showed the converse pattern: absolute error was substantially lower, but geographic correlation was weaker, and the dispersion of predicted prevalence varied from close agreement with observations in Hawaiʻi to marked expansion in New York State. Thus, a synthetic population may approximate the average prevalence of a health behavior without reproducing its geographic distribution, or preserve relative differences between communities while remaining poorly calibrated in absolute terms.^14^ These distinctions are consequential because different applications of population simulation, from identifying geographic disparities to estimating population burden or evaluating interventions, may depend on different dimensions of fidelity.

The distribution of prediction error further identified a systematic limitation of the current framework. Across both Hawaiʻi and New York State, overestimation was greatest among communities with the lowest observed screening prevalence, particularly for colorectal cancer screening, and diminished as observed prevalence increased. This pattern suggests that LLMPopSim more readily reproduced central tendencies than the extremes of the population distribution. Such attenuation is especially important in a public health context, where communities at the extremes of behavioral or disease distributions may be those for which accurate characterization is most consequential.^15^ A population simulator that systematically shifts low prevalence communities toward the population mean could obscure meaningful geographic disparities even when overall error or correlation appears acceptable. Preserving the tails of population distributions should therefore be considered an important criterion for evaluating generative population models, particularly if these systems are ultimately intended to model heterogeneous responses to public health interventions.

Performance across community characteristics was heterogeneous rather than following a uniform socioeconomic gradient. In New York State, colorectal cancer screening error increased most clearly across increasing uninsured prevalence and, to a lesser extent, poverty and population size, whereas mammography showed different and often nonmonotonic patterns. Similarly, in Hawaiʻi, colorectal cancer screening error was relatively stable across income, poverty, educational attainment and population size, although error was higher in ZCTAs with greater Native Hawaiian and Pacific Islander representation and increasing uninsured prevalence. These patterns did not extend uniformly to mammography, for which error instead varied most prominently with population size and uninsured prevalence. Together, these findings suggest that population simulation error may depend on interactions among the behavior being modeled, geography and community context rather than reflecting a single axis of socioeconomic disadvantage. Evaluating performance across population subgroups and community characteristics will therefore be important as generative population models are extended to more diverse settings, particularly to ensure that aggregate performance does not obscure systematic limitations in the communities they are intended to represent.^8,16^

The potential value of this approach extends beyond small-area estimation. Conventional small-area models are designed to estimate population-level outcomes directly from observed data,^1,2,6,7^ whereas LLMPopSim represents those outcomes as the aggregate behavior of individually characterized synthetic agents. This distinction creates the possibility of moving from prediction toward simulation: rather than estimating only what is observed in a population, future generative population models could allow characteristics of individuals, communities or interventions to be modified and their resulting population-level effects examined. Recent agent-based frameworks illustrate this potential: AgentTorch integrates LLM-driven behavior into large-scale population simulations supporting retrospective, prospective and counterfactual analyses, while VacSim uses demographically grounded generative agents to simulate responses to vaccine-related public health interventions.^12,17^ Such applications could provide a computational environment for exploring how heterogeneous populations may respond to changes in screening access, eligibility, health messaging or other public health interventions. The present study does not establish the validity of LLMPopSim for such counterfactual applications; rather, small-area estimation provides an empirically measurable setting in which to test whether simulated individual behavior aggregates into realistic population-level patterns before more complex uses are considered. Establishing this correspondence represents an important step toward synthetic populations that can ultimately serve not only as representations of observed communities, but as experimentally manipulable models of population health.

Several limitations define important priorities for further development. First, the synthetic populations were constructed from area-level marginal distributions and simplified demographic assumptions, including fixed age and sex distributions, and therefore did not fully preserve the joint relationships among demographic, socioeconomic and health characteristics that exist within real populations. More realistic population synthesis incorporating empirically derived joint distributions may improve both individual-level plausibility and aggregate geographic fidelity.^14^ Second, LLMPopSim incorporated a ZCTA-level screening prior derived from community characteristics, making the resulting estimates a product of both community-level contextualization and individual-agent simulation. Future work should disentangle the relative contributions of these components and determine how much population-level structure emerges from individual agents themselves. Third, validation was limited to two preventive health behaviors, two states and a single LLM, and the observed differences between colorectal cancer screening and mammography demonstrate that performance for one behavior cannot be assumed to generalize to another. Finally, CDC PLACES estimates provide a practical population-level benchmark but are themselves modeled estimates rather than direct measurements of individual behavior.2 Broader validation across health behaviors, geographic settings, model architectures and independently observed population outcomes will therefore be necessary before generative population models can be used reliably for counterfactual or policy simulation.

Overall, this study provides an initial empirical foundation for geographically grounded LLM-based population simulation. By demonstrating that the decisions of individually represented synthetic agents can aggregate into community-level patterns that retain measurable features of real-world health behavior, and that these patterns persist, albeit imperfectly, across temporal and geographic transfer, LLMPopSim extends generative agent modeling from the simulation of individuals toward the simulation of populations. At the same time, the observed differences in calibration, geographic correlation, distributional fidelity and subgroup performance demonstrate that realistic population simulation requires substantially more than reproducing an average outcome. Continued advances in population synthesis, behavioral modeling and external validation will be necessary before such systems can support credible intervention or policy experiments. Ultimately, generative population models could provide a complementary approach for studying how heterogeneous individuals and communities respond to changing health conditions and interventions, creating experimentally tractable synthetic populations for questions that are difficult to examine directly in real-world populations.

## Methods

### I. Study Design

We developed LLMPopSim, a large language model (LLM)-based generative population simulation framework designed to estimate community-level preventive health behaviors. Framework development and individual agent prompt optimization were performed using historical Hawaiʻi cohorts from 2018 and 2020. Following optimization, the finalized framework was fixed and evaluated without further prompt modification using 2022 Hawaiʻi data for temporal evaluation and 2022 New York State data for geographic evaluation. Primary outcomes included ZCTA-level colorectal cancer screening and mammography prevalence. Framework performance was evaluated using measures of predictive accuracy, calibration, preservation of geographic variability, and subgroup performance.

#### Data Sources

Publicly available data from the U.S. Census Bureau American Community Survey (ACS) 5-year estimates were used to characterize demographic and socioeconomic features of each ZCTA. Core ACS-derived variables included total population, median household income, proportion below the federal poverty threshold, educational attainment, unemployment, and mean household size. For the Hawaiʻi cohorts, the proportion of residents identifying as Native Hawaiian or Pacific Islander was additionally derived for state-specific subgroup analyses. This variable was not provided to the LLM during either ZCTA-level prevalence estimation or individual-agent behavioral simulation. Healthcare access was represented using the corresponding CDC PLACES measure rather than an ACS-derived insurance variable.

Community-level health characteristics and preventive health behavior estimates were obtained from CDC PLACES. PLACES-derived measures encompassed healthcare access, chronic disease, cardiometabolic risk factors, health behaviors, preventive healthcare utilization, oral health, and self-reported health status. The ACCESS2 measure was used to represent lack of health insurance among adults aged 18-64 years. PLACES releases were selected according to the underlying data year corresponding to each study cohort: the 2020 PLACES release was used for the 2018 Hawaiʻi development cohort, the 2022 PLACES release for the 2020 Hawaiʻi development cohort, and the 2024 PLACES release for the 2022 Hawaiʻi and New York State evaluation cohorts. ZCTA-level crude prevalence estimates were extracted and merged with ACS-derived demographic and socioeconomic variables by ZCTA to generate geographically resolved feature distributions. Preventive health outcomes evaluated in this study included colorectal cancer screening (COLON_SCREEN) and mammography (MAMMOUSE).

#### Study Cohorts

Framework development and individual agent prompt optimization were performed using Hawaiʻi cohorts from 2018 (93 ZCTAs) and 2020 (92 ZCTAs). Following prompt optimization, the finalized framework was evaluated using independent 2022 cohorts from Hawaiʻi (94 ZCTAs) and New York State (1,745 ZCTAs) to assess temporal and geographic generalizability, respectively. ZCTAs were included when corresponding ACS and CDC PLACES data could be successfully linked and the demographic, socioeconomic, health-related, and outcome fields required for the corresponding analysis were populated. ZCTAs with missing required data were excluded from the relevant analysis; these exclusions occurred predominantly among ZCTAs with small populations.

### II. Synthetic Population Generation

#### Construction of Joint Distributions of Features

For each study year and geographic setting, demographic and socioeconomic variables from the ACS were merged with community-level health measures from CDC PLACES by ZCTA to construct a Joint Distribution of Features (JDoF). Each JDoF comprised a ZCTA-level feature profile containing the population-level parameters used for synthetic population generation, including demographic and socioeconomic characteristics and crude prevalence estimates for selected health-related measures. These parameters were subsequently used to define the probability distributions from which individual synthetic agent attributes were sampled. Preventive health behaviors serving as prediction targets were excluded from the features used to generate agents for the corresponding behavioral simulation.

#### Generation of Synthetic Individuals

Synthetic adult populations were generated independently for each ZCTA using probabilistic sampling informed by the corresponding ZCTA-level feature profile. The number of synthetic adults generated for each ZCTA was defined as 77% of the corresponding total ACS population, reflecting an approximate adult population share applied uniformly across ZCTAs, states, and study years. Outcome-specific age and sex eligibility criteria were subsequently applied to these synthetic populations to identify agents eligible for colorectal cancer screening or mammography polling. Age and sex were generated from fixed population-level distributions that were held constant across ZCTAs, states, and study years. Age was sampled from a normal distribution with a mean of 45 years and standard deviation of 15 years, truncated to ages 18-90 years, and sex was sampled using probabilities of 51% female and 49% male. Individual income was sampled from a log-normal distribution centered on the corresponding ZCTA median household income, with a log-scale standard deviation of 0.6. Household size was sampled from a normal distribution centered on the corresponding ZCTA mean household size, with a standard deviation of 0.5 and a lower bound of one person. Educational attainment was sampled from three categories, high school or less, bachelor’s degree, and graduate degree, using ZCTA-specific proportions for bachelor’s and graduate degrees, with the remaining probability assigned to high school or less. Health-related characteristics derived from CDC PLACES were assigned as binary agent attributes using Bernoulli sampling according to the corresponding ZCTA-level crude prevalence estimates. These included measures of healthcare access, chronic disease, cardiometabolic risk factors, health behaviors, preventive care, oral health, and self-reported health. The screening outcome serving as the prediction target was excluded from agent features to prevent direct outcome leakage; the alternate screening behavior was retained as a non-target contextual feature. Eligibility for behavioral polling was defined according to the age and sex criteria corresponding to each preventive health measure. For colorectal cancer screening, eligible agents were aged 50-75 years in the 2018 and 2020 cohorts and 45-75 years in the 2022 cohorts, reflecting the change in screening eligibility over the study period. For mammography, eligible agents were female and aged 50-74 years across all study cohorts. Synthetic population generation was performed using a fixed random seed of 123.

#### Agent Representation

Synthetic individuals meeting the eligibility criteria for the target screening behavior were converted into structured agent profiles for LLM polling. Each profile contained age, sex, household income, household size, educational attainment, ZCTA, and available non-target health-related attributes derived from CDC PLACES. PLACES-derived attributes included healthcare access, chronic conditions, cardiometabolic risk factors, health behaviors, preventive care, oral health, and self-reported health measures. The screening outcome being predicted was excluded from the agent representation to prevent direct outcome leakage; the alternate screening behavior was retained as a non-target contextual feature.

### III. LLM-Based Behavioral Simulation

#### LLM Inference

Llama 3.3 70B Chat was used for all LLM inference throughout the study, including ZCTA-level prevalence prior estimation and individual-agent behavioral polling across all study cohorts and preventive health outcomes. Inference was performed on the Oermann laboratory high-performance computing environment using an internally hosted OpenAI-compatible endpoint.

#### ZCTA-Level Prevalence Prior Estimation

Before agent-level behavioral simulation, an initial ZCTA-level prevalence estimate was generated for each target preventive health behavior. For each ZCTA, the model received a structured community-level profile comprising population size and socioeconomic characteristics derived from the ACS, including household income, poverty, educational attainment, unemployment, and household size, together with non-target community health characteristics derived from CDC PLACES. The screening outcome being estimated was excluded from the profile to prevent direct outcome leakage, whereas the alternate screening behavior was retained as a non-target contextual feature. Race/ethnicity variables used for subsequent subgroup analyses were not provided to the LLM. A single LLM inference was performed per ZCTA for each outcome at a temperature of 0.0, yielding an estimated percentage of eligible adults who were up to date with the corresponding screening behavior. No repeated sampling or averaging of ZCTA-level estimates was performed. This estimate served as a fixed prevalence prior for all subsequent individual agent polling within that ZCTA for the corresponding outcome.

#### Behavioral Polling

Synthetic individuals meeting the eligibility criteria for each target preventive health behavior were submitted to the LLM in batches of 10 agents per request, with the model returning one binary behavioral prediction for each agent in the batch. For each request, the model received the structured profiles of the included agents together with the corresponding ZCTA-level prevalence estimate generated during the preceding community-level estimation step. The model was instructed to treat this estimate as a prior prevalence when assigning individual binary decisions indicating whether each agent was up to date with the target screening behavior. Agent-level decisions were subsequently aggregated within each ZCTA, with simulated prevalence calculated as the mean of binary predictions among eligible agents. LLM inference was performed with a temperature of 0 and a fixed inference seed of 123. All eligible agents within each ZCTA were polled without subsampling. Agent order was deterministically shuffled using a fixed seed of 42 before polling. Model outputs were constrained to structured JSON format and parsed to extract the binary screening decision for each agent.

#### Prompt Development and Optimization

Prompt optimization was restricted to the individual-agent behavioral polling stage; the prompt used to generate the initial ZCTA-level prevalence estimates was not optimized. Individual-agent prompt development was performed jointly using the 2018 and 2020 Hawaiʻi development cohorts. Beginning with an initial prompt, candidate revisions were iteratively generated and evaluated by repeating agent-level polling on the development cohorts. Candidate performance was assessed using a composite objective designed to balance absolute predictive accuracy, geographic discrimination, and preservation of geographic variability. The objective was defined as MAE + 0.03(1 – r) + 0.05|1 – SD ratio|, where r represents the Pearson correlation between predicted and observed ZCTA-level prevalence and SD ratio represents the ratio of the standard deviation of predicted prevalence to that of observed prevalence. Lower objective values indicated better performance. RMSE, mean prediction bias, calibration slope and intercept, and predicted-to-observed interquartile range ratios were additionally calculated as diagnostic measures but were not included in the optimization objective. Candidate prompts were accepted when the composite objective improved by at least 0.005, and optimization was terminated after two consecutive iterations without improvement meeting this threshold. Optimization was conducted independently for colorectal cancer screening and mammography using the same procedure; both optimization processes converged on the same final prompt. The resulting prompt was fixed before evaluation on the 2022 Hawaiʻi and New York State test cohorts.

### IV. Evaluation Framework

#### Primary Performance Measures

LLMPopSim-predicted ZCTA-level prevalence estimates were compared with observed CDC PLACES prevalence estimates for colorectal cancer screening and mammography. Mean absolute error (MAE) was prespecified as the primary measure of predictive accuracy and was calculated as the mean absolute difference between predicted and observed ZCTA-level prevalence. Complementary performance measures included root mean squared error (RMSE), mean prediction bias, and Pearson correlation coefficient (r). Together, these metrics characterized absolute prediction error, systematic over- or underestimation, and the ability of the framework to preserve relative differences in screening prevalence across ZCTAs. Performance metrics were calculated separately for each combination of study cohort and preventive health outcome. In the 2022 test datasets, colorectal cancer screening and mammography were therefore evaluated independently in Hawaiʻi and New York State, yielding four state-outcome evaluations. No further prompt modification or model calibration was performed using the 2022 test data.

#### Calibration and Geographic Variability

Calibration was assessed separately for each study cohort and preventive health outcome by fitting a linear regression of observed CDC PLACES prevalence on LLMPopSim-predicted prevalence across ZCTAs. Calibration slope and intercept were estimated from these models, with a slope of 1 and intercept of 0 representing ideal calibration. Agreement between predicted and observed prevalence was additionally evaluated using scatterplots and residual analyses. Preservation of geographic variability was assessed by comparing the standard deviation of predicted prevalence with that of observed prevalence across ZCTAs. The predicted-to-observed standard deviation ratio (SD ratio) was used as the primary measure of variability preservation, with a value of 1 indicating equivalent geographic dispersion, values below 1 indicating attenuation of geographic variability, and values above 1 indicating greater variability in predicted than observed prevalence.

#### Subgroup and Error Analyses

To evaluate whether prediction performance varied across community characteristics, ZCTAs in both 2022 test cohorts were stratified into quartiles according to five demographic and socioeconomic characteristics available across states: population size, median household income, proportion of residents below the poverty threshold, proportion with a bachelor’s degree, and PLACES-estimated proportion uninsured among adults aged 18-64. For the Hawaiʻi cohort, additional analyses stratified ZCTAs according to the proportion of Native Hawaiian and Pacific Islander (NHPI) residents. Mean absolute error and mean prediction bias were calculated within each quartile separately for colorectal cancer screening and mammography. To characterize error across the outcome distribution, ZCTAs were additionally stratified into quartiles of observed screening prevalence, and mean residual error and absolute error were calculated within each quartile. Finally, associations between continuous ZCTA characteristics and prediction error were explored using Pearson correlation coefficients. These analyses were intended to characterize patterns of model performance across community features rather than to establish independent causal or inferential associations.

#### Statistical Analysis

Final statistical analyses were performed in R version 4.5.1 (R Foundation for Statistical Computing). Mean absolute error, RMSE, mean prediction bias, Pearson correlation coefficients, calibration slope and intercept, and predicted-to-observed standard deviation ratios were calculated separately for each study cohort and preventive health outcome. Calibration was assessed using linear regression of observed prevalence on predicted prevalence. Exploratory associations between continuous ZCTA characteristics and prediction error were assessed using Pearson correlation coefficients. Subgroup analyses were descriptive and compared prediction error across quartiles of demographic and socioeconomic characteristics and observed screening prevalence. All analyses were conducted at the ZCTA level.

## Data Availability

All source data used in this study are publicly available. Demographic and socioeconomic data were obtained from the U.S. Census Bureau American Community Survey (ACS) 5-year estimates (2018, 2020, and 2022), available at https://www.census.gov/programs-surveys/acs/data.html. Community-level health characteristics and preventive health behavior estimates were obtained from the Centers for Disease Control and Prevention (CDC) PLACES project (2020, 2022, and 2024 releases), available at https://www.cdc.gov/places/tools/data-portal.html. Individual-level populations used for behavioral simulation were synthetically generated from these publicly available aggregate data as described in the manuscript.

https://www.cdc.gov/places/tools/data-portal.html

## Author Contributions

M.N. and E.K.O. conceived and designed the study. M.N. developed the methodology, performed the analyses and simulations, interpreted the results, generated the figures, and drafted the manuscript. R.J.S. and X.H. provided computational infrastructure and technical support. E.K.O. supervised the study. All authors reviewed and approved the manuscript.

## References

1. Pierannunzi C, Xu F, Wallace RC, et al. A methodological approach to small area estimation for the Behavioral Risk Factor Surveillance System. Prev Chronic Dis. 2016;13:E91. doi:10.5888/pcd13.150480.

2. Greenlund KJ, Lu H, Wang Y, et al. PLACES: local data for better health. Prev Chronic Dis. 2022;19:210459. doi:10.5888/pcd19.210459.

3. Doubeni CA, Selby K, Gupta S. Framework and strategies to eliminate disparities in colorectal cancer screening outcomes. Annu Rev Med. 2021;72:383–398. doi:10.1146/annurev-med-051619-035840.

4. Ahmed AT, Welch BT, Brinjikji W, et al. Racial Disparities in Screening Mammography in the United States: A Systematic Review and Meta-analysis. J Am Coll Radiol. 2017;14(2):157-165.e9. doi:10.1016/j.jacr.2016.07.034.

5. Khan-Gates JA, Ersek JL, Eberth JM, Adams SA, Pruitt SL. Geographic Access to Mammography and Its Relationship to Breast Cancer Screening and Stage at Diagnosis: A Systematic Review. Womens Health Issues. 2015;25(5):482–493. doi:10.1016/j.whi.2015.05.010.

6. Zhang X, Holt JB, Yun S, et al. Validation of multilevel regression and poststratification methodology for small area estimation of health indicators from the Behavioral Risk Factor Surveillance System. Am J Epidemiol. 2015;182(2):127–137. doi:10.1093/aje/kwv002.

7. Wang Y, Holt JB, Xu F, et al. Using 3 health surveys to compare multilevel models for small area estimation for chronic diseases and health behaviors. Prev Chronic Dis. 2018;15:E133. doi:10.5888/pcd15.180313.

8. Wesson P, Hswen Y, Valdes G, Stojanovski K, Handley MA. Risks and opportunities to ensure equity in the application of big data research in public health. Annu Rev Public Health. 2022;43:59–78. doi:10.1146/annurev-publhealth-051920-110928.

9. Aiello AE, Renson A, Zivich PN. Social media- and internet-based disease surveillance for public health. Annu Rev Public Health. 2020;41:101–118. doi:10.1146/annurev-publhealth-040119-094402.

10. Park JS, O’Brien J, Cai CJ, Morris MR, Liang P, Bernstein MS. Generative agents: interactive simulacra of human behavior. In: UIST ‘23: Proceedings of the 36th Annual ACM Symposium on User Interface Software and Technology. Association for Computing Machinery; 2023:Article 2, 1–22. doi:10.1145/3586183.3606763.

11. Park JS, Zou CQ, Kamphorst J, et al. LLM agents grounded in self-reports enable general-purpose simulation of individuals. Preprint. Posted online November 15, 2024. Updated June 28, 2026. arXiv:2411.10109 [cs.AI]. doi:10.48550/arXiv.2411.10109.

12. Hou AB, D. H, Wang Y, et al. Can a society of generative agents simulate human behavior and inform public health policy? A case study on vaccine hesitancy. Preprint. Posted online March 12, 2025. Updated July 13, 2025. arXiv:2503.09639 [cs.MA]. doi:10.48550/arXiv.2503.09639. Accepted to COLM 2025.

13. Liu R, Jong C, Li H, et al. Simulating population compliance with pandemic interventions using large language models. Preprint. Posted online May 15, 2026. medRxiv 2026.05.12.26352942. doi:10.64898/2026.05.12.26352942.

14. Chapuis K, Taillandier P, Drogoul A. Generation of synthetic populations in social simulations: a review of methods and practices. J Artif Soc Soc Simul. 2022;25(2):6. doi:10.18564/jasss.4762.

15. Shah SN, Russo ET, Earl TR, Kuo T. Measuring and monitoring progress toward health equity: local challenges for public health. Prev Chronic Dis. 2014;11:E159. Published 2014 Sep 18. doi:10.5888/pcd11.130440.

16. Qu Y, Wang J. Performance and biases of Large Language Models in public opinion simulation. Humanit Soc Sci Commun. 2024;11:1095. doi:10.1057/s41599-024-03609-x.

17. Chopra A, Kumar S, Giray-Kuru N, Raskar R, Quera-Bofarull A. On the limits of agency in agent-based models. Preprint. Posted online September 14, 2024. Updated November 10, 2024. arXiv:2409.10568 [cs.MA]. doi:10.48550/arXiv.2409.10568.

